# Investigation of Research Quality and Transparency in Neurosurgery Through the Utilization of Open Science Practices

**DOI:** 10.1101/2024.08.03.24311452

**Authors:** Zahin Alam, Kush Desai, Anirudh Maddali, Vijay Sivan, Rohit Prem Kumar, Geoffrey R. O’Malley, Nitesh Patel

**Affiliations:** Department of Neurosurgery, Hackensack Meridian School of Medicine, Nutley, NJ; Department of Neurosurgery, HMH-Jersey Shore University Medical Center, Neptune, NJ

**Keywords:** Neurosurgical journals, open science practices, research quality, research transparency, research reproducibility

## Abstract

**Background and Objective:** Neurosurgical research is a rapidly evolving field, with numerous studies continuously published. As the body of research grows, upholding high-quality standards becomes increasingly essential. Open science practices offer tools to ensure quality and transparency. However, the prevalence of these practices remains unclear. This study investigated the extent to which neurosurgical publications have implemented open science practices.

**Methods:** Five open science practices (preprint, equator guidelines, published peer review comments, preregistration, and open accessibility to data and methods) were measured from five top-ranked neurosurgical journals (*Neurosurgery, Journal of Neurosurgery, World Neurosurgery, Neurosurgical Review*, and *Acta Neurochirurgica*), according to Google Scholar. One hundred fifty articles were randomly sampled from January 1, 2022, to December 31, 2023. Two reviewers analyzed these articles for their utilization of open science practices. A third reviewer settled disagreements.

**Results:** One journal required (20%) and three journals (60%) recommended utilizing EQUATOR guidelines. Three journals (60%) allowed preprints, and all five journals (100%) recommended or required preregistration of clinical trials, but only two (40%) recommended preregistration for systematic reviews (Figure 1). All five journals (100%) recommended or required methods to be publicly available, but none (0%) published peer-review comments. *Neurosurgical Review* utilized the most open science practices, with a mean utilization of 1.4 open science practices per publication versus 0.9 across the other four journals (p < 0.001). Moreover, *Neurosurgical Review* significantly utilized more open science practices versus *Journal of Neurosurgery* (p < .05) and *World Neurosurgery* (p < .05). Both randomized controlled trials (p < .001) and systematic reviews (p < .001) significantly utilized more open science practices compared to observational studies.

**Conclusions:** Despite advocacy from neurosurgical journals, the adoption of open science practices still needs improvement. Implementing incentives and clearer requirements may prove beneficial. Promoting these practices is crucial to enhancing transparency and research quality in neurosurgery.

## INTRODUCTION

Open science practices refer to scientific research’s reproducibility and transparency, including open data, methods, results, and accessibility. These have increasingly become a part of research in the medical field, and efforts have been made to encourage open practices in all scientific research. In 2023, the Biden-Harris administration, through the White House Office of Science and Technology, launched the *Year of Open Science*, which included actions “to advance national open science policy, provide access to the results of the nation’s taxpayer-supported research, accelerate discovery and innovation, promote public trust, and drive more equitable outcomes.” ^1^ Lack of transparency in research has various implications, such as giving providers and patients incomplete information about the risks and benefits of treatments. Furthermore, it can lead to unnecessary spending of vital health system funds on ineffective therapies or guidelines.^2,3^ Open science practices enable evidence-based and transparent decisions in medicine, which bolsters trust among patients and researchers.^1,3,4^

Inconsistency in research practices has also resulted in the reproducibility crisis, where published research findings often cannot be replicated. Moreover, research waste, including biases and methodological errors, has been estimated to account for 85% of all medical research funding.^4,5^ Lievore et. al. found an inverse relationship between the percentage of publications retracted and the Journal Citation Report (JCR).^6^ This indicated that errors in publications and lack of reproducibility led to fewer citations and use of those publications, hindering scientific progress and contributing to overall mismanagement. Between 1992 and 2012, papers retracted due to misconduct alone accounted for about $58 million of NIH research funding.^7^ As a result, there is a critical need for improved regulation of research quality, which open science standards aim to address. However, it is essential to evaluate whether these standards are being properly implemented and working, particularly across various types of journals. Research practices have been investigated diffusely throughout medicine, but there is no current literature on the use of open science practices in the neurosurgical literature. Thus, the purpose of this study was to explore the use of these open-science practices in papers published in top neurosurgical journals.

## METHODS

### Cohort Selection

The top five neurosurgical journals, according to Google Scholar, were selected after the exclusion of affiliate journals such as *Neurosurgical Focus* and *Journal of Neurosurgery: Spine*. The five journals selected were *Neurosurgery, Journal of Neurosurgery, World Neurosurgery, Neurosurgical Review, and Acta Neurochirurgica*. Articles between January 1, 2022, and December 31, 2023 were gathered. One hundred fifty articles, 30 articles from each journal, were randomly sampled and selected with the use of a random number generator.

### Data Collection

Five key open science variables were collected: 1) the availability of preprints, 2) adherence to EQUATOR guidelines, 3) publication of peer review comments, 4) preregistration of studies, and 5) open access to data and methods. One reviewer independently investigated journal guidelines for each of the five journals. The objective of this particular review was to ascertain whether each journal offered guidelines for their authors that required, suggested, or failed to mention each of the five open science practices.

Focusing on the article review, two reviewers independently analyzed each publication and evaluated the adherence of each study to the five open science practices. Furthermore, each article was classified by its study type (randomized controlled trial, observational study, or systematic review). Availability of preprints was evaluated by whether the preprint was accessible on a preprint server or if the preprint was cited in the background or methods. Articles were considered to have followed EQUATOR guidelines if the proper guidelines (PRISMA, CONSORT, STROBE, etc.) were mentioned in the manuscript. Preregistration was determined by study type. Randomized controlled trials (RCTs) were classified as preregistered if the date their study protocol was listed on clinicaltrials.gov or a similar international registry before the commencement of the study. Observational studies (OS) were considered preregistered if their study protocol was registered in a database such as protocols.io, PROSPERO, researchregistry.com, or osf.io. Systematic reviews (SRs) were categorized as preregistered if their study protocol was registered in PROSPERO. Articles were considered to have published peer-review comments if these comments were explicitly mentioned in the manuscript. Articles that fulfilled the criteria for open access to data and methods were assessed based on their supplementary material. This criterion was considered met if there was publication of raw data, code, expanded study methods, or if a data availability statement was included. Any disagreements between the two independent reviewers were reconciled by the senior author.

### Statistical Analysis

Comparison testing between journals and study types was conducted by ANOVA with pairwise comparisons. Groups of journals were compared to each other using a *t*-test. Linear regression determined if there was an association between journal rank and open science practice use. All analyses were done utilizing Microsoft Excel (Version 2405, Microsoft Corporation, Redmond, Washington). P-values < 0.05 were considered significant.

## RESULTS

One journal required (20%), and three journals (60%) recommended utilizing EQUATOR guidelines. Three journals (60%) allowed preprints, and all five journals (100%) recommended or required preregistration of clinical trials, but only two (40%) recommended preregistration for systematic reviews. All five journals (100%) recommended or required methods to be publicly available, but none (0%) published peer-review comments (Figure 1).

**Figure 1.**
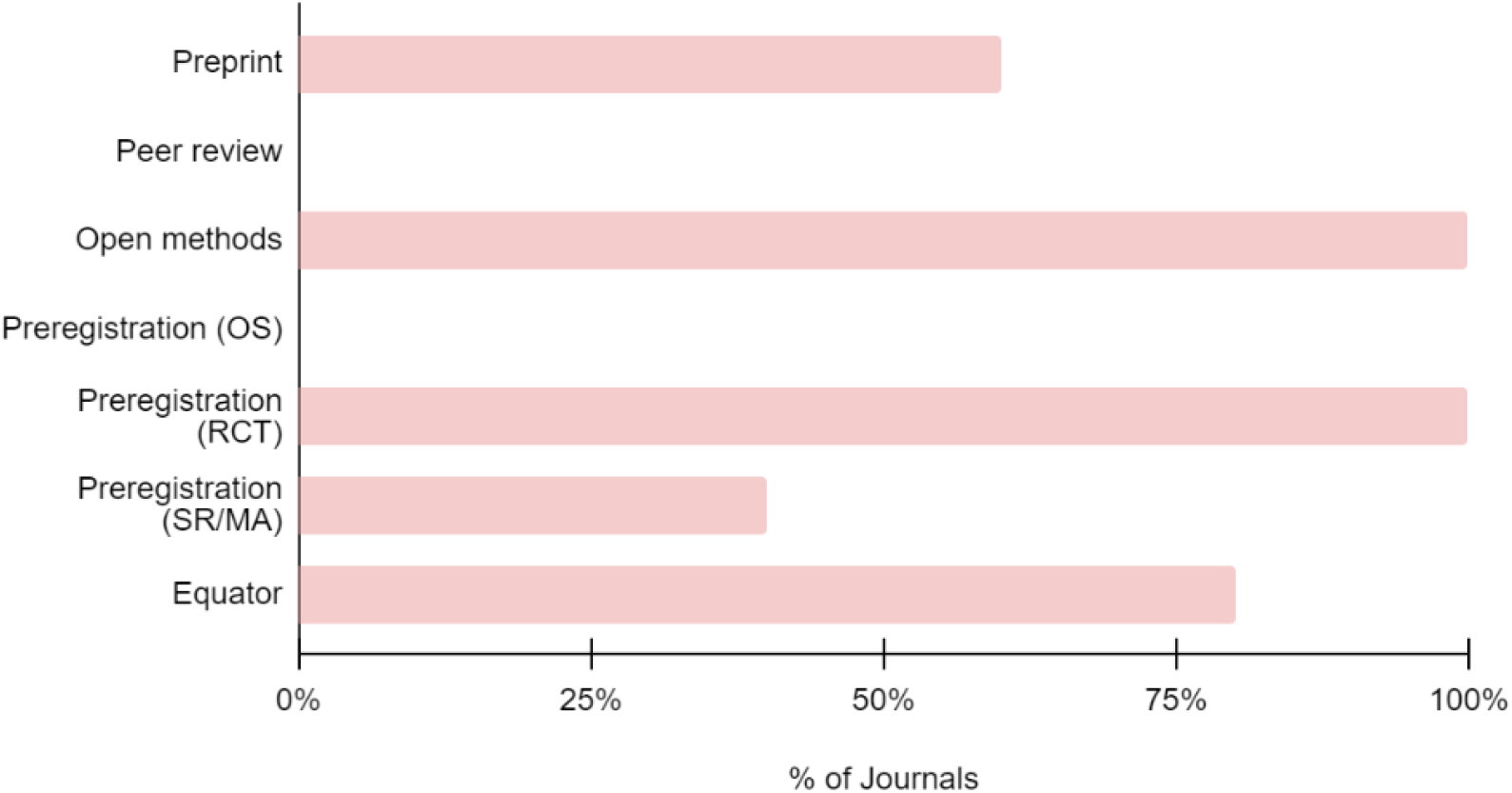
Percentage of neurosurgical journals (*n=5*) in recommending or requiring each open science practice within their journal guidelines.

Across all 150 studies, 109 (73%) utilized at least one or more open science practices. Ninety-four studies (63%) from our sample properly reported EQUATOR guidelines. Twenty-four articles (16%) provided open access to data, while no studies published preprints (0%) or peer review comments (0%). Preregistration rates varied by study type, with all RCTs (100%) preregistered. OSs (6%) and SRs (17%) provided less adherence to preregistration (Figure 2). Throughout these 150 articles, only 9 (6%) studies required reconciliation by the third reviewer.

**Figure 2.**
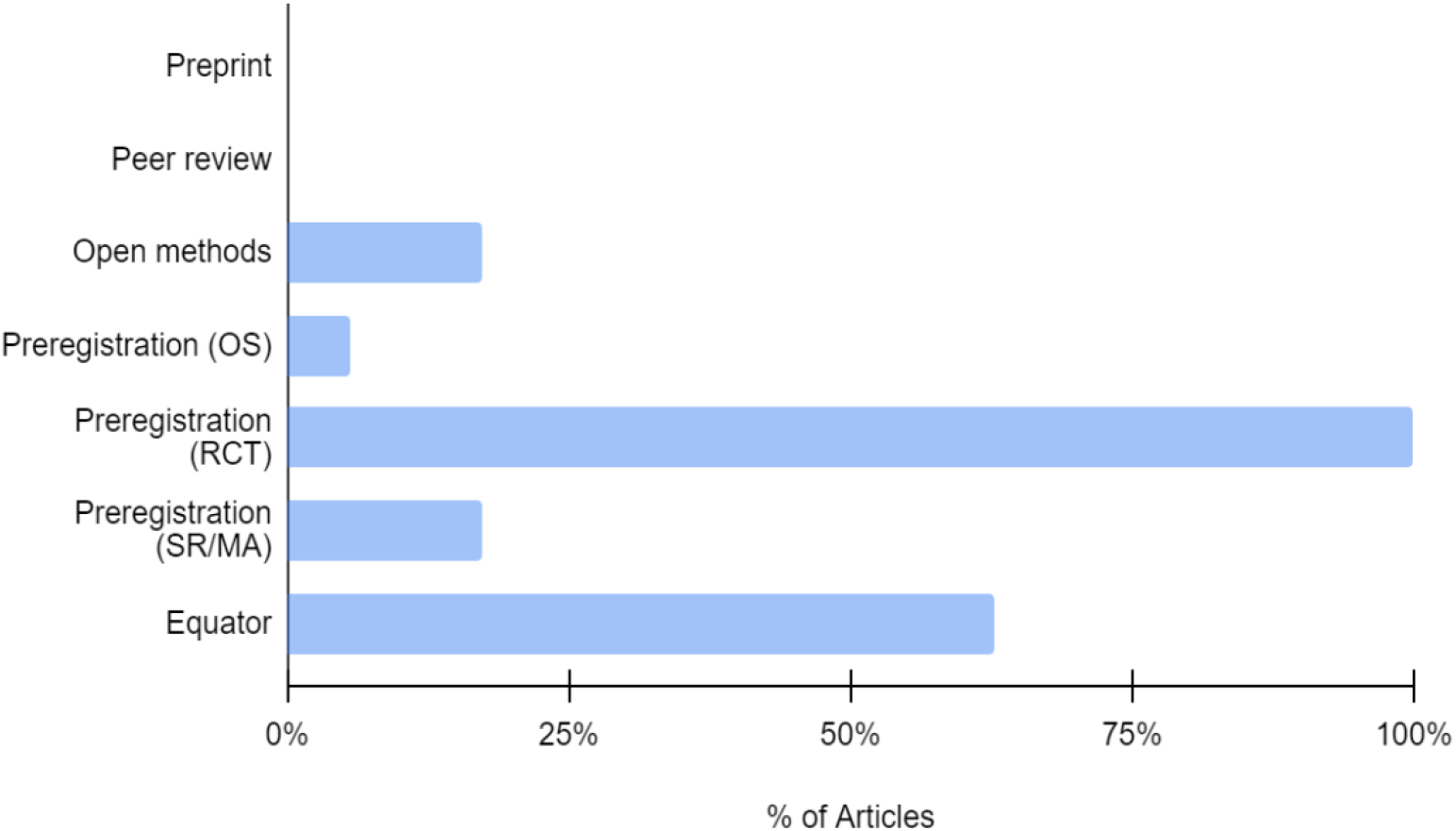
Percentage of open science utilization among all 150 articles.

*Neurosurgical Review* utilized the most open science practices, with a mean utilization of 1.4 open science practices per publication versus 0.9 across the other four journals (p < 0.001). Moreover, *Neurosurgical Review* significantly utilized more open science practices versus the *Journal of Neurosurgery* (p < 0.050) and *World Neurosurgery* (p < 0.050). There was no relationship between journal rank and the use of open science practices (R^2^ = 0.32)

RCTs and SRs utilized the most open science practices per publication, with a mean utilization of 1.2 and 1.3 open science practices, respectively. In contrast, OSs utilized the least open science practices with a mean utilization of 0.3 practices per publication. Both RCTs (p < 0.001) and SRs (p < 0.001) significantly utilized more open science practices compared to OSs. There was no significant difference in open science practice utilization between RCTs and SRs (p = 0.916).

## DISCUSSION

This study investigated the utilization of open science practices in five leading neurosurgical journals. Findings revealed that there is significant variability in their application of open science usage in both the journal and article levels. One journal required and three journals recommended the use of EQUATOR guidelines. Notably, all five journals recommended or required methods to be publicly available, but none published peer-review comments. *Neurosurgical Review* demonstrated the highest mean utilization of open science practices per publication. When analyzing the utilization of open science practices in specific types of studies, RCTs and SRs applied the most open science practices per publication.

Despite the efforts of journals to implement open science practices, adherence to these practices remains generally poor. The use of open science practices in imaging journals found that 65% had policies on protocol registration and reporting guidelines.^8^ However, similar to our results in neurosurgical journals, adherence at the article level was significantly lower and variable, potentially highlighting a disconnect between editorial policy and open science practice implementation. Nonetheless, journal policies could benefit from further refinement. For example, policies regarding data availability in general and internal medicine journals are often inconsistent, and this inconsistency could potentially lead to confusion among researchers seeking to adhere to these guidelines.^9^ Additionally, Grant et. al analyzed the implementation of ten Transparency and Openness Promotion (TOP) standards within a broad range of journals (339) and highlighted that while some journals implement various TOP standards, most lack a comprehensive adherence to open science practices or consistent enforcement at the article level.^10^ Moreover, multiple smaller studies have demonstrated that various types of journals exhibit low compliance with transparency guidelines.^11–15^ Specifically, general surgery journals were poorly compliant with specific open science guidelines. Of the 240 articles they surveyed, 34% used one or more open science practices, 26% complied with EQUATOR guidelines, 5% pre-registered protocols, and 9% fully disclosed methods to the public.^16^ The low use of open science standards in neurosurgical journals, as found in our study, aligns with several of these aforementioned findings, demonstrating that publicly available practices and guidelines may be insufficient to achieve a truly transparent research community. Hence, there is a necessary call to action for several journal types, including neurosurgical journals, to better promote open science usage and to prevent unintended consequences. For example, non-adherence to open science practices can delay advances in treatment and management guidelines, as illustrated by previous research on post-heart-transplant antibody-mediated-rejection (AMR). Scientific research in the 1990s found evidence that AMR most often presented in the first three months post-transplant and would often recur within the first year post-transplant.^17^ Further, patients with more than 3 AMR episodes were likely to die of cardiovascular-related causes. However, it took 24 years since the first description of post-heart transplant AMR for diagnostic criteria to be published in official guidelines. Despite having the knowledge, the development of guidelines was delayed by affinity bias and publication bias.^17^ Open science guidelines aim to address these concerns by collaborating knowledge and thereby accelerating the translation of research findings into clinical applications.

The quality and reproducibility of studies published in neurosurgical journals have previously been investigated, revealing potential gaps and areas for improvement. Among 633 review articles across ten leading specialty neurosurgical journals, only 45.97% of search strategies were reproducible.^18^ Rothoerl et al. explored the level of evidence (LOE) and citation index across neurosurgical journals. 55% of 982 articles investigated were rankable by LOE, and only 0.4% and 3.0% of these studies met the criteria for LOE Ia. and LOE Ib, respectively. The mean citation index across all 982 articles was suboptimal at 4.80, although for literature reviews, clinical studies, and experimental studies, the citation indices were 10.66, 6.78, and 5.03, respectively.^19^ Therefore, it is crucial to advocate for increased adoption of open science practices, as these measures can significantly enhance neurosurgical research. However, there may be several specific challenges to applying open science practices in surgical journals diffusely.

Research transparency promotion, such as through data-sharing policies, is quite limited in surgical journals.^20–22^ Therefore, many surgical studies, specifically concerning RCTs, have not been reproducible.^23^ These results could allude to the fact that implementing open science practices in surgical research could be quite difficult due to unique challenges. These challenges may stem from the complex nature of surgical research, which involves balancing patient safety, privacy, and intellectual property with the need for transparency and reproducibility. However, these challenges should be addressed, as incomplete transparency in methodology can significantly impact research conclusions. In a recent study examining outcomes for head and neck reconstruction surgery using the National Surgical Quality Improvement Program (NSQIP) database, different approaches to case selection and statistical analysis yielded significantly varying results, thereby highlighting the need for transparency when reporting methods.^24^ This allows other researchers to reproduce the work published in surgical journals while also aiding publishing journals in evaluating methodologies and improving surgical literature aimed at maximizing surgical outcomes.

Additionally, other barriers to promoting open science usage include a lack of institutional funding and incentives, the presence of fees in adhering to open science practice, and the lack of mandates. Thus, incentives and “badges” can be implemented to recognize publications and journals that adopt open science practices, which would highlight their commitment to research publication quality control and encourage others to integrate these practices in their work. The Development Psychology Department of Brigham Young University introduced an incentivized quality measurement system for evaluating faculty research. The measurement system grades research quality based on criteria such as the use of preregistration and adherence to EQUATOR guidelines. The purpose of this system was to encourage better science practice and reduce pressure on faculty to accumulate publications and novel findings, and instead focus on valuable but underutilized research initiatives that implement longitudinal studies, larger sample sizes, and other time-consuming criteria.^25,26^ The hope is that this system motivates other faculties and institutions to adopt similar incentivized initiatives and address challenges seen between the competitive nature of academia and open sharing.^27^ Moreover, many have expressed uncertainty and ambiguity about the exact requirements needed for them for open science usage.^27–29^ The numerous guidelines and expectations can cause confusion among researchers and lead to improper usage. In a survey of 198 cardiology researchers, most participants indicated that they received no formal training with open science practices and utilized these strategies at their discretion.^30^ To enhance journal communication with researchers, it is essential to include specific and standardized instructions that detail how to improve transparency in their submissions. Journal requirements have been shown to influence researcher’s ensuing practices.^31^ This approach would ensure clarity and consistency in expectations regarding transparency practices.

Despite these barriers, there is growing sentiment and motivation for the scientific community to expand open science usage for the advancement of scientific knowledge.^32–34^ Researchers were surveyed on outstanding motivators for open science practices. For personal motivation, open science was mentioned to potentially improve researchers’ professional standing. Open access would encourage unanticipated collaboration between groups, creating a more diverse and visible community and augmenting publication productivity. Furthermore, due to the competitive nature of scientific research, open science could be used strategically to maintain strong research metrics. For example, data sharing with other groups could prove useful if proper attribution is included, such as through citation, acknowledgment, or co-authorship. From a broader perspective, interviewees highlighted ethical motivation as open sharing would help reduce delays in the publication of publicly funded research and prevent “bottlenecking” of data prior to publication. Broadly sharing data samples would foster greater access to research output and maximize research efficiency and discovery.^34^

### Limitations

This study is subject to certain limitations. Due to the review of a sample of 150 publications, there may be a biased estimation of open science usage. A more thorough, extensive investigation that involves a larger selection of journals and articles is required. Additionally, while we utilized several preprint and preregistration databases, it’s possible that a preprint or preregistration was registered in other locations.. Lastly, EQUATOR guidelines may not have been mentioned within the manuscript, and there is a possibility that certain articles followed the guidelines but did not state them.

## CONCLUSION

This study aims to pave the way for the analysis of the adherence of journals to open science protocols. There is ample room for further development in this subject. For example, similar studies should be conducted in other surgical subspecialties. Other studies can continue to investigate the validity and potential of open science practices.^35^ By promoting the incorporation of these open science protocols, a high standard of quality throughout the research community can be maintained. Improved research quality can directly lead to better patient outcomes, enhanced patient safety measures, and stronger trust between the provider and patient with reliable research results and conclusions.

## Data Availability

All data produced in the present study are available upon reasonable request to the authors

## Data availability Statement

The datasets generated and/or analyzed during the current study are available from the corresponding author upon reasonable request.

